# Increased brain coverage and efficiency when measuring current-induced magnetic fields by use of simultaneous multi-slice echo-planar MRI

**DOI:** 10.1101/2025.06.21.25329642

**Authors:** Teresa Cunha, Fróði Gregersen, Lars G. Hanson, Axel Thielscher

## Abstract

**Purpose:** Magnetic resonance current density imaging (MRCDI) can non-invasively validate electric field simulations in volume conductor head models. Weak electric currents are injected using scalp electrodes while measuring the MR phase perturbations caused by the tiny magnetic fields (1-2 nT) induced by the current flow in tissue. MRCDI generally has a low signal-to-noise ratio, making it susceptible to technical imperfections and physiological noise. Here, we tested and optimized simultaneous multi-slice (SMS) EPI for time-efficient and robust brain MRCDI.

**Methods:** MRCDI data was acquired in a phantom and five human brains using SMS-EPI optimized for measuring current-induced phase perturbations. Multiband factors and interslice gaps were systematically varied and the resulting image quality assessed. In particular, the impact of interslice signal leakage on the measured phase was tested.

**Results:** Current-free acquisitions showed the expected noise amplification with decreasing interslice distances. However, physiological noise generally dominated the human data, making the overall noise levels identical to single-slice EPI for interslice gaps of at least 12 mm and multiband factors between 3 and 5. Upon application of electric currents, the phantom data revealed subtle artifacts for multiband factors 5 and 6, even for large gaps. Nevertheless, artifacts were absent in the human brain for multiband factors up to 5, where the performance of SMS-EPI approached that of single-slice measurements for sufficient interslice distances.

**Conclusion:** Optimized SMS-EPI with multiband factors up to 5 and minimum interslice gaps of 12 mm performs on par with single-slice EPI, making it attractive for increasing brain coverage in MRCDI.

## 1. Introduction

Stimulation targeting and dose control in transcranial brain stimulation can benefit from electric field simulations in personalized volume conductor models of the head[1]. Invasive measurements in patients and non-human primates have demonstrated reasonable fits between the simulated and measured fields on average, but also strong deviations in single cases, suggesting the need for improvements[2], [3], [4]. Magnetic resonance current density imaging (MRCDI) is a promising non-invasive technique for validating and optimizing electric field simulations. Injection of weak electric currents using scalp electrodes is combined with MRI to reconstruct current density distributions[5]. These currents induce a magnetic flux density distribution that locally changes the precession frequency of the spins in the sample. Consequently, the phase of the measured signal is modulated by ΔB_z,c_, the component of the current-induced magnetic field that is parallel to the main field B_O_. In brain MRCDI in-vivo, the weak electric currents that can safely and comfortably be injected (1-2 mA)[6], [7], [8] induce magnetic fields of only a few nT. Recently, we optimized a double-echo gradient echo EPI sequence with short repetition time for single-slice imaging of the small ΔB_z,c_-induced phase modulations. We demonstrated that it achieves a good sensitivity while being robust to physiological noise due to its high temporal resolution[9]. However, the relatively long scanning time required to measure ΔB_z,c_ with a sufficient signal-to-noise ratio (SNR) can prevent the acquisition of multiple slices to achieve beneficial[10] whole-brain coverage in practical brain MRCDI experiments.

Here, we optimized and validated simultaneous multi-slice (SMS) imaging for a time-efficient increase of brain coverage in EPI-based MRCDI. Unlike standard in-plane parallel imaging which shortens the acquisition time by reducing the number of phase-encoding (PE) steps for each slice, k-space can be fully sampled in SMS acquisitions to minimize SNR losses and reduce the “g-factor” penalty[11]. However, increasing the number of simultaneously acquired slices (i.e. the multiband - MB - factor) can intensify the mixing of signal between slices, visible as so-called “leakage” artifacts[12], [13]. Their severity has been vastly mitigated in SMS-EPI by introducing in-plane shifts between the simultaneously acquired slices (“blipped-CAIPI”) and reconstructing using the Slice-GRAPPA method[14], where each slice is separated by convolving the SMS k-space data with the corresponding GRAPPA-like kernels (estimated from a low-resolution single-slice reference scan). The Split Slice-GRAPPA method further reduces the leakage artifacts by trading off the total error during the kernel fitting procedure against the amount of interslice leakage[15].

While SMS-EPI has become a mainstay of functional and diffusion MRI, it has not yet been validated and optimized for MRCDI[16] that is particularly challenging due to its need for highly accurate MR phase data. Guided by our prior studies[7], [10] we chose the settings for SMS-EPI that maximize its sensitivity to ΔB_z,c_ while maintaining image quality. Moreover, we optimized the analyses to account for the effects of B_O_ inhomogeneity on the ΔB_z,c_ images. We then evaluated the quality of the magnetic fields measured with different MB factors and interslice distances, in both phantom and human brain experiments. Although the results also depend on the coil and sequence characteristics, they give insight into the relevant SMS parameter choices for high-quality ΔB_z,c_ measurements. This study also describes procedures and provides reference values for optimizing and evaluating local measurement protocols.

## 2. Methods

### 2.1. Data acquisition

Scanning was performed in a 3T MRI scanner (MAGNETOM Prisma, Siemens Healthcare, Erlangen, Germany) using a 64-channel head coil and the “Multi-Band EPI C2P” sequence (https://www.cmrr.umn.edu/multiband/) developed by the Center for Magnetic Resonance Research (CMRR, Minneapolis, Minnesota, USA). It implements the blipped-CAIPI approach for a more efficient slice separation, with relative in-plane shifts of 1/3 of the field of view (FOV) along the PE direction automatically imposed for MB factors > 2 and no in-plane acceleration. Additionally, it offers two methods for optimizing the slice-separation kernels: the default Split Slice-GRAPPA (“LeakBlock”)[15] and the Slice-GRAPPA[14]. Unless otherwise stated, the former was used for disentangling the slices.

Two k-space traversals followed each excitation pulse, allowing for a higher bandwidth along the PE direction to reduce geometric distortions caused by B_0_ inhomogeneities[17], while maintaining a long data acquisition period to increase SNR. All experiments were performed with echo times T_E_ = 25.6, 63.5 ms, repetition time T_R_ = 120 ms, flip-angle 30°, matrix size 64 x 64 and voxel dimensions 3.4 x 3.4 x 3 mm^3^, in accordance with our prior parameter optimizations[7], [10]. Each multiband acquisition was time-matched to that of one single slice.

Acquisitions without electric currents were initially performed to assess the noise level in the ΔB_z,c_ measurements (denoted “noise floors”). Measurements with applied currents were then performed using a “loop setup”, where the currents are not injected into the sample but flow through a cable placed around it instead (Figures 2B&3C). This allows for rigorous validation of the measured ΔB_z,c_, since the magnetic fields from the cable currents can be calculated from the Biot-Savart integrals[18]. Electric currents with an intensity of 2 mA baseline-to-peak were applied, inverting the polarity at the beginning of each repetition of the EPI sequence.

Lastly, a 3D ultra-short T_E_ PETRA[19] structural scan was acquired in each scanning session to image the conductive rubber cable[20] and then simulate ΔB_z,c_ using the Biot-Savart law. The simulations were then subtracted from the measurements with currents to assess their accuracy.

### 2.2. ΔB_z,c_ computations

ΔB_z,c_ was computed as the difference in the phase images from consecutive repetitions of the double-echo EPI sequence, divided by the phase sensitivity of the sequence to the current-induced magnetic fields (2γT_E_, with γ denoting the proton gyromagnetic ratio)[10]. The ΔB_z,c_ images from each echo time were then combined based on their variance, as estimated from the temporal SNR of the magnitude images at the respective echo times[21]. Regions where the ΔB_z,c_ measurements were not considered trustworthy were masked out as described in Supporting Material S1. The ΔB_z,c_ images were finally corrected for geometric distortions caused by B_0_ inhomogeneities using field maps estimated from the phase images at the two echo times, as explained in Supporting Material S2.

### 2.3. Phantom experiments

Experiments were performed on a spherical FBIRN phantom with a diameter of 17 cm and relaxation times comparable to human brain tissue (T1/T2 ≈ 530/50 ms).

Initially, the “LeakBlock” and Slice-GRAPPA kernels were tested with a MB factor of 6 and a small interslice gap of 6 mm to provoke leakage artifacts. Measurements with and without electric currents were performed, each including 1000 repetitions of the EPI sequence. Piloting showed that, despite the lower noise levels, the Slice-GRAPPA method resulted in severe leakage artifacts in the ΔB_z,c_ measurements with electric currents (Figure S2). Thus, all experiments described below used the “LeakBlock” kernels for slice separation.

In an experiment without currents, the MB factors {3, 4, 5, 6} and interslice gaps {6, 9, 12, 15, 18} mm were tested (the gap being the nominal distance from the upper to the lower edge of consecutive slices). The slices associated with different MB factors were organized as depicted in Figure 1A, ensuring that at least three slices were directly comparable across MB factors. When varying the interslice distance, one of the slices was kept fixed, making measurements comparable across the different gaps (slice s2, dashed green line in Figures 1B&C). Each measurement included 200 repetitions of the EPI sequence.

**Figure 1.**
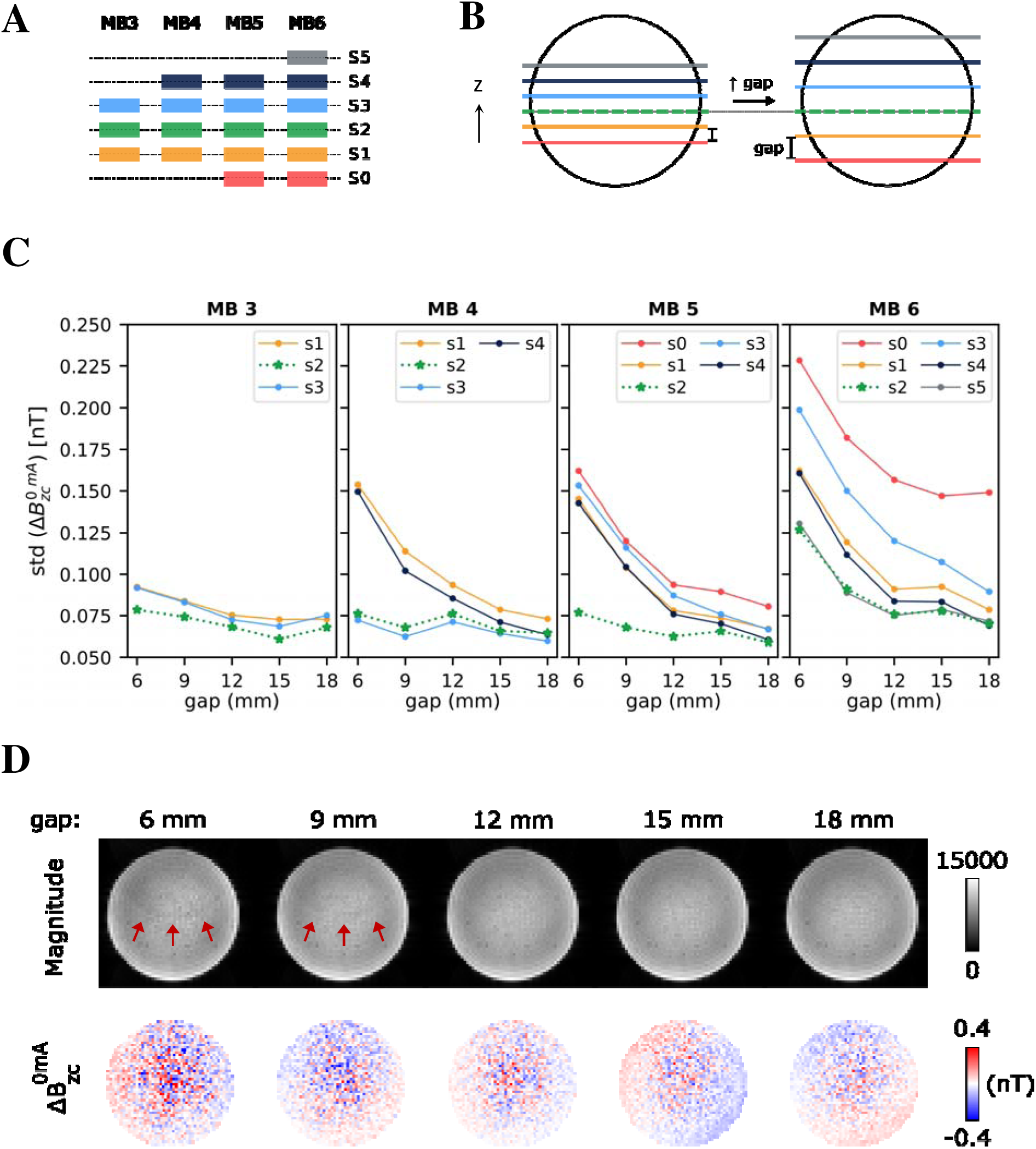
Adopted strategy and results from the ΔB_z,c_ measurements without current injection in a phantom. **A)** Relative positioning of the slices (s0 to s5) across multiband (MB) factors. **B)** Slice shifting along *z* with increasing interslice gap. Note that the position of slice number 2 (s2, dashed green line) was kept fixed. **C)** Summary of the noise levels in the ΔB_z,c_ measurements, as estimated from the spatial standard deviation of the ΔB_z,c_ images obtained without current injection, for the different MB factors and interslice gaps. **D)** Magnitude and ΔB_z,c_ images of the fixed slice (s2) obtained with a MB factor of 6 and different interslice gaps. Subtle artifacts are noticeable on the magnitude images for the smallest gaps and are indicated by red arrows.

**Figure 2.**
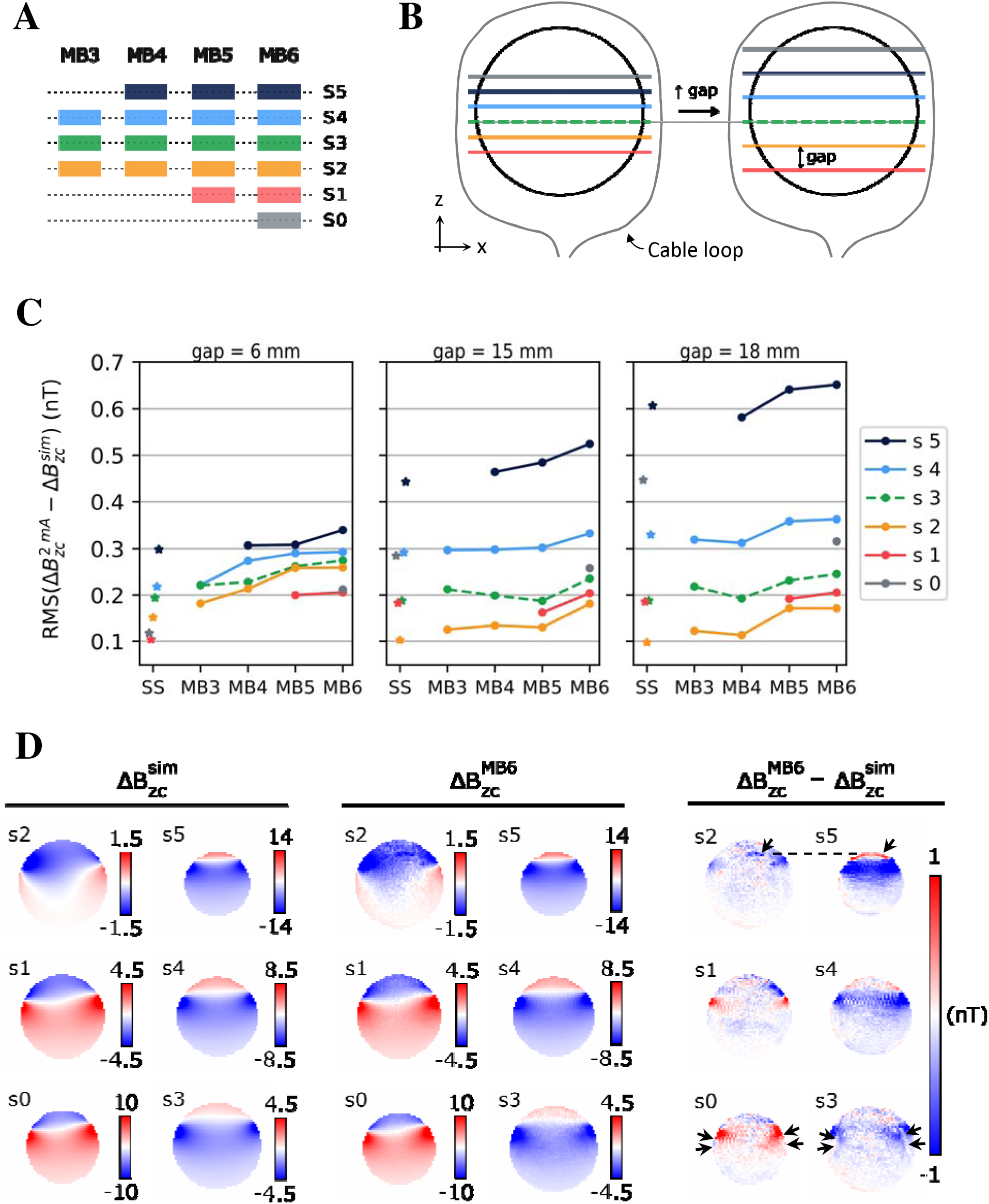
Adopted strategy and results from the ΔB_z,c_ measurements with current loops around a phantom. **A)** Relative positioning of the slices (s0 to s5) across multiband (MB) factors. **B)** Loop setup and slice shifting along *z* with increasing interslice gaps. **C)** Root Mean Square (RMS) error between the measured (ΔB_z,c_^2mA^) and simulated (ΔB_z,c_^sim^) current-induced magnetic fields. Results are shown for multi-slice measurements with different MB factors and interslice gaps, and the corresponding single-slice (SS) measurements. Please note that as the interslice gap increased, the outermost slices got closer to the lead, where the current-induced magnetic fields were stronger, also causing increased RMS errors. **D)** Example of the simulated and measured current-induced magnetic fields, and respective differences for an acquisition with MB factor 6 and interslice gap of 18 mm. Slices sharing the same CAIPI-shift are shown side by side. The black arrows in s2 and s3 highlight the presence of subtle artifacts, likely due to signal leaking from s5 and s0, respectively.

Based on the results from the current-free experiment, a reduced parameter space including the MB factors {3, 4, 5, 6} and the interslice gaps {6, 15, 18} mm was tested in a new experiment with electric currents (Figure 2A-C). The exclusion of the interslice distances 9 and 12 mm was based on the noisier results (compared to MB 3) obtained for the other MB factors with gaps smaller than 15 mm. A 6 mm gap was still included as the expected worst-case scenario. Finally, validated single-slice measurements were performed at all positions sampled by the SMS acquisitions, each consisting of 500 repetitions of the EPI sequence.

### 2.4. Human experiments

Five healthy volunteers with no MRI contraindications were included in this part of the study after written informed consent. The study complied with the Helsinki Declaration on human experimentation and was approved by the Ethics Committee of the Capital Region of Denmark (De Videnskabsetisk Medicinske Komitéer, approval number 2206924).

Given the results from the phantom experiments, MB factor 6 was not further tested in humans. However, all interslice gaps between 6 and 18 mm were again included as additional noise sources contaminating human data might make the relative contributions of the leakage artifacts less relevant. This resulted in the following parameter space: MB = {3, 4, 5} and gap = {6, 9, 12, 15, 18} mm (Figure 3A-D). Single slices were imaged at all positions sampled by the SMS acquisitions, each including 500 repetitions of the EPI sequence.

**Figure 3.**
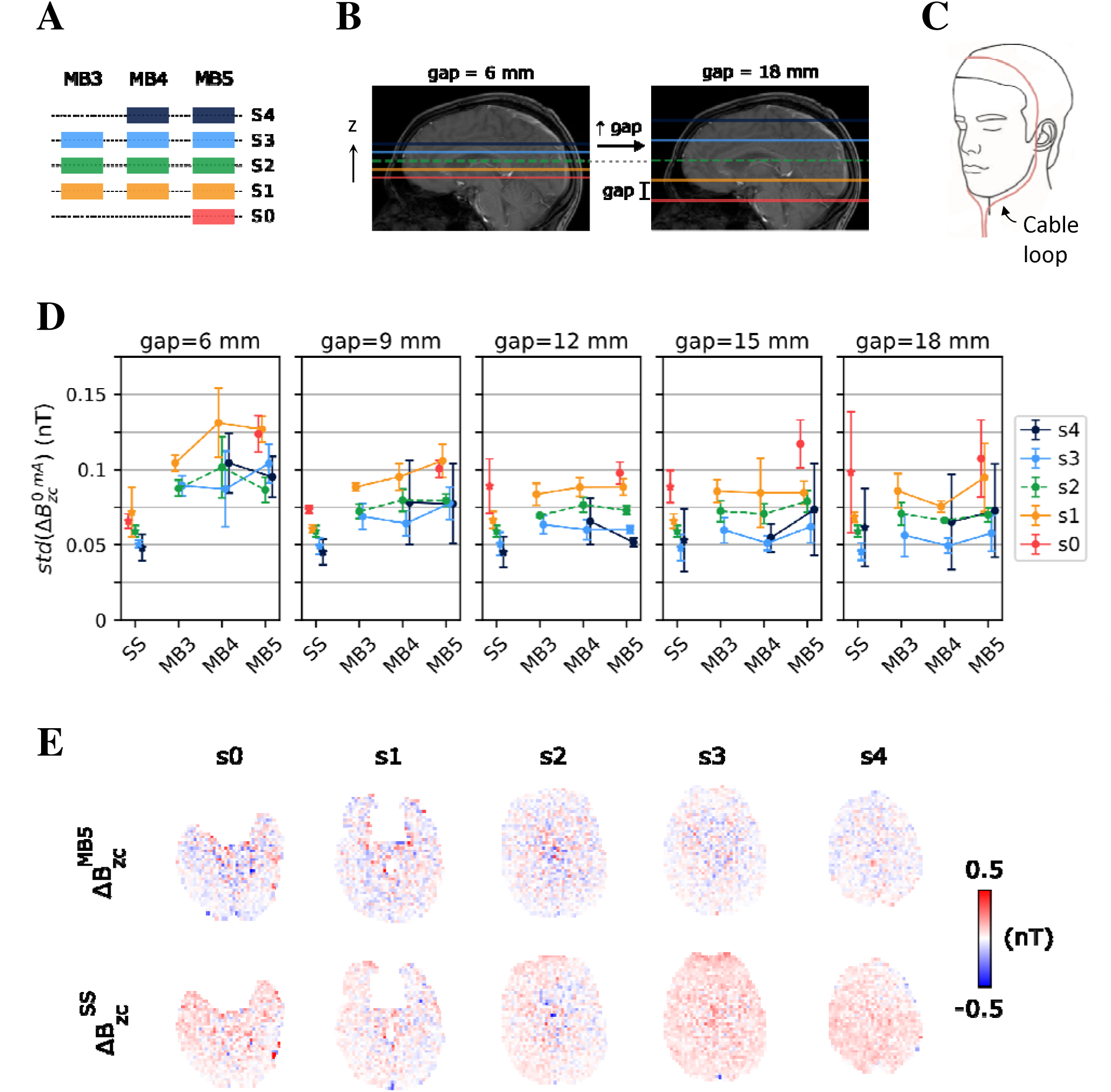
Adopted strategy and results from the ΔB_z,c_ measurements without current injection in the human brain. **A)** Relative positioning of the slices (s0 to s4) across multiband (MB) factors. **B)** Slice shifting along z with the increase in the interslice gap. Note the significant change in brain coverage from the smallest to the largest gap. **C)** Loop setup (the corresponding results are shown in Figure 4). **D)** Summary of the noise levels in the ΔB_z,c_ measurements, as estimated from the spatial standard deviation of the ΔB_z,c_ images obtained without current injection. Results are shown for multi-slice measurements with different MB factors and interslice gaps, and the corresponding single-slice (SS) measurements. Each data point and vertical error bar represent the average and standard deviation across five subjects. **E)** ΔB_z,c_ noise floor images from an SMS acquisition with a MB factor of 5 and an interslice gap of 12 mm (first row), and corresponding single-slice acquisitions (second row).

## 3. Results

### 3.1. Phantom experiments

Figure 1C summarizes the noise floor measurements, where the noise was quantified as the spatial standard deviation of the current-free ΔB_Z,C_ images. Slice s2 was kept in a fixed position, thus being suited for evaluating the effects of varying interslice gaps. As expected, the noise levels decreased as the gap increased. This was particularly noticeable for MB 6, suggesting that challenging SMS data benefits most from the larger variation in the coil sensitivities along *z* attained with larger gaps. Examples of magnitude and ΔB_Z,C_ noise floor images of slice s2 are shown in Figure 1D for MB 6. Subtle artifacts are seen in the magnitude images for the smallest interslice distances. The previously mentioned attenuation of the random noise in ΔB_Z,C_ with increasing gap is clearly visible.

Since the absolute positions of the remaining slices changed with the interslice distance, the MB factors should be compared for each specific gap (Figure 1C). The biggest differences occurred for the smallest gaps: MB 3 resulted in relatively low noise levels in all slices, whereas only two, one and no slices achieved equally low noise with MB factors 4-6, respectively. This pattern is consistent with the apparent noise “coupling” between pairs of slices noticeable for MB factors > 3, which is likely related to the imposed CAIPI shifts. For example, slice 1 showed higher noise levels for MBs 4-6 than for MB 3, and its noise behavior mirrored that of slice 4. Slice 4 was absent for MB 3 and otherwise shared the same in-plane CAIPI shift as slice 1, which likely increases the signal leakage between them. Increasing the gaps to ≥15 mm helped to achieve equally low noise levels in MB factors 3-5, while higher levels still occurred in some slices for MB 6.

The performance of single-slice and SMS acquisitions upon application of electric currents is summarized in Figure 2C. Shown are the root mean square (RMS) errors between the measured ΔB_Z,C_ and the simulations based on the Biot-Savart Law. In general, higher MB factors tended to result in slightly larger errors, although this trend was weak. In particular, MB factors 3-4 consistently showed equally good performance, comparable to single-slice acquisitions. MB factor 5 with an 18 mm gap and MB 6 with 15/18 mm gaps showed slightly increased noise levels. For those conditions, subtle artifacts occurred in some slices, potentially caused by signal leaking between slices sharing the same CAIPI shift (the black arrows in Figure 2D indicate examples where leakage between slices 2 and 5, and between slices 0 and 3 seem to occur). Please note that, due to the chosen cable configuration, the magnetic fields increase along the *z* direction. Thus, the increase of the absolute RMS errors for the higher slices (most obvious for slice s5) does not indicate a lower measurement quality at those positions, but rather reflects that the chosen error metric scales with the ΔB_Z,C_ strength.

### 3.2. Human Experiment

SMS showed generally worse SNR over the time series of magnitude images than single-slice acquisitions. This often resulted in the exclusion of slightly larger brain regions during the first step of the masking procedure described in Supporting Material S1. However, taking single-slice measurements as reference, the number of voxels in the masks stayed clearly above 90% even for MB 5 (on average across subjects; Figure S3). In the current-free experiment, a low number of acquisitions were affected by measurement instabilities of unclear origin. Therefore, noise floor images with RMS values exceeding the average by two standard deviations were considered outliers, resulting in the exclusion of 3 out of 95 single-slice and 2 out of 75 SMS acquisitions.

The noise levels stayed below 0.2 nT in all the remaining measurements (Figure 3D). SMS achieved larger noise levels than single-slice acquisitions, mostly for the smallest gap (6 mm), while this effect quickly decreased for larger interslice distances. All three tested MB factors performed similarly. In Figure 3E, a representative example of the noise floor images obtained with a MB factor of 5 and a 12 mm gap is given, along with the corresponding single-slice measurements. The accuracy of the ΔB_z,c_ measurements with currents in the cable loop was also assessed through comparison with the Biot-Savart simulations (Figure 4A). The RMS error (RMSE) between measurements and simulations did not differ substantially between the three MB factors tested and were similar to the single-slice acquisitions. RMSEs below 0.3 nT were obtained for most measurements, with higher errors observed for the top slice (s4) especially for the largest gaps, which placed it in higher brain regions. Figure 4B shows representative results with a MB factor of 5 and a 12 mm gap, along with the corresponding single-slice measurements. There was good agreement between the ΔB_z,c_ measured with the two acquisition types and the simulations, with no severe penalty resulting from the high acceleration factor.

**Figure 4.**
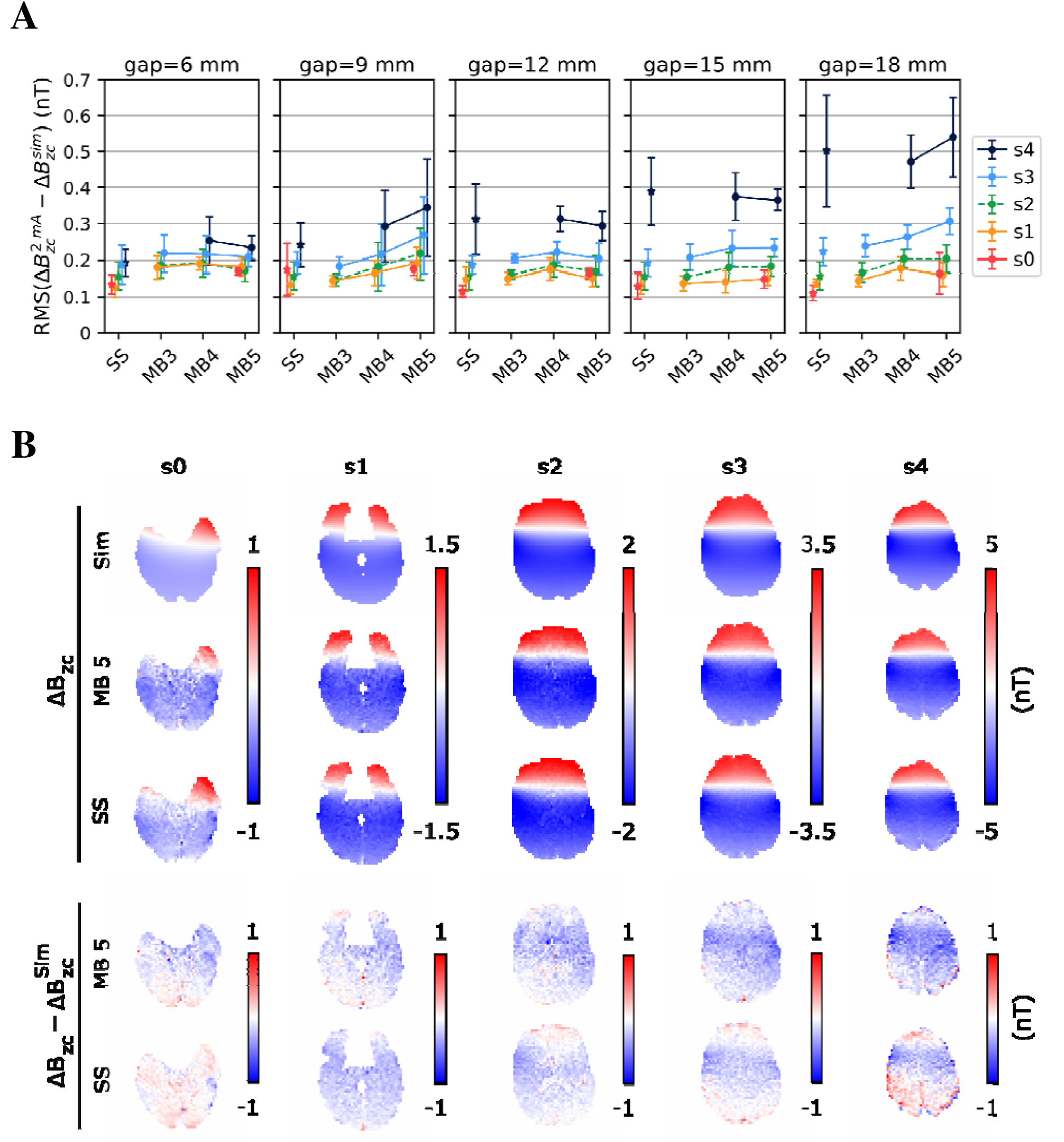
Results from the ΔB_z,c_ measurements with current injection in the human brain. The slices (s0 to s4) were positioned as shown in Figure 3A. **A)** Root Mean Square (RMS) error between the measured (ΔB_z,c_^2mA^) and simulated (ΔB_z,c_^sim^) current-induced magnetic fields. Results are shown for multi-slice measurements with different multiband (MB) factors and interslice gaps, and the corresponding single-slice (SS) measurements. Each data point and vertical error bar represent the average and standard deviation across subjects. **B)** Comparison between a multi-slice acquisition with a MB factor of 5 and an interslice gap of 12 mm, and corresponding single-slice acquisitions. Top: simulated and measured magnetic fields. Bottom: differences between the measured and simulated fields.

## 4. Discussion

In this study, we optimized and validated SMS-EPI acquisitions of current-induced magnetic fields for human brain MRCDI. Initial phantom tests without current flow confirmed the expected increase of the noise levels with decreasing interslice gaps and increasing MB factors, which both make the separation of signals from different slices more difficult[22], [23]. Moreover, closer inspection of the noise floors highlighted the importance of the in-plane CAIPI shifts for reducing interslice leakage. For example, with MB 3, the imposed shifts (in multiples of FOV_PE_ /3) ensured that all slices were shifted in relation to each other. In contrast, with MB 4, the top and bottom slices shared the same shift, making their separation harder and increasing noise floors in both slices. Importantly, however, the relevance of interslice leakage decreased with increasing interslice gaps, and MBs 4 and 5 reached similar noise levels to MB 3 for gaps of 12 mm or more.

In subsequent phantom tests with current flow in a cable loop, SMS revealed error levels (compared to simulations) that remained very similar to those of single-slice acquisitions across the tested parameter space.

The current-free human brain measurements also showed noise levels comparable to those of single-slice acquisitions for all tested MB factors with interslice gaps of 12 mm or more. Accordingly, a combination of MB 5 with a 12 mm gap appears as a promising setting for SMS-EPI-based brain MRCDI, providing good brain coverage and maintaining low noise levels.

In the measurements with current flow, ΔB_z,c_ increased along the *z* direction due to the chosen cable configuration both in the phantom and human experiments. As our error metric relies on absolute differences between measurements and simulations, the increasing error levels with increasing interslice gaps observed in the higher slices (most obvious for slices s4 and s5 in Figures 2 and 4) are thus expected. Since good quality noise floors were measured in those slices, we believe that the observed errors do not reflect a lower measurement quality but are mainly caused by inaccuracies in the Biot-Savart simulations due to imprecise cable tracking, in line with our previous observations[18].

Recently, deep learning-based SMS reconstruction methods have shown comparable [24] or even better [25] performance than traditional SENSE- or GRAPPA-like approaches. However, these new methods have mainly been validated on magnitude data, and rigorous assessment of their performance on phase data is still missing. It is outside the scope of this paper to evaluate these newly emerging slice-separation techniques for use with MRCDI. The focus of this study was to extend our most recent EPI-based MRCDI method [9] to SMS-EPI using an easily accessible and well tested sequence (CMRR “Multi-Band EPI C2P” sequence). We hope that this will also benefit the broader usage of MRCDI in the future.

SMS imaging is generally combined with in-plane acceleration to achieve higher acceleration factors [23], [26], [27] at the cost of reduced SNR and increased “g-factor” penalty. In-plane acceleration is, however, not very suitable for EPI-based MRCDI. In fact, reducing the duration of each EPI readout will not substantially reduce the total acquisition time of each ΔB_z,c_ measurement, as the echo times (and consequently, repetition time) need to be long enough to allow for sufficient current-induced phase accumulation.

A remaining limitation of our EPI-based MRCDI method (both for SMS and single-slice) is the inefficient spoiling of signal from long T2* tissues, leading to the exclusion of parts of the cerebrospinal fluid (CSF) during the masking procedure. Improving spoiling to obtain usable data from CSF would require increasing the spoiler gradient strengths[6], which was hindered by the lack of access to the source code of the sequence. Furthermore, flow sensitivity would remain.

## 5. Conclusion

SMS-EPI provided good measurements of the current-induced magnetic fields that were mostly on par with single-slice results, therefore offering an attractive time-efficient way of increasing brain coverage in MRCDI. Within the tested parameter space, only the smallest interslice gap of 6 mm exhibited higher noise floors in humans compared to single-slice acquisitions, while the remaining combinations of MB factors and gaps performed identically.

Our findings suggest flexibility in the choice of parameters based on the desired brain coverage. For example, combining a moderate interslice distance (e.g., 12 mm) with MB factors no larger than 5 will vastly improve the brain coverage compared to single-slice EPI, while maintaining a very similar quality of the measured magnetic fields. While measurement quality is also influenced by the specific coil and sequence implementation used, our study generally confirms the advantages of SMS-EPI for highly sensitive measurements in brain MRCDI.

## Data Availability

Data of the human subjects cannot be shared publicly because of privacy restrictions. The phantom data is fully available on OSF (DOI 10.17605/OSF.IO/DCWUN, https://osf.io/dcwun/).

## 6. Acknowledgments

This study was supported by the Lundbeck Foundation (https://lundbeckfonden.com/, grant R313-2019-622 to AT, grant R324-2019-1784 to LGH) and the German Research Foundation (https://www.dfg.de/, DFG grants TH 1330/6-1 and TH 1330/7-1, part of Research Unit FOR 5429 “MeMoSLAP”). The funders had no role in study design, data collection and analysis, decision to publish, or preparation of the manuscript.

## Supplementary Material

**Supplementary Figure S1.**
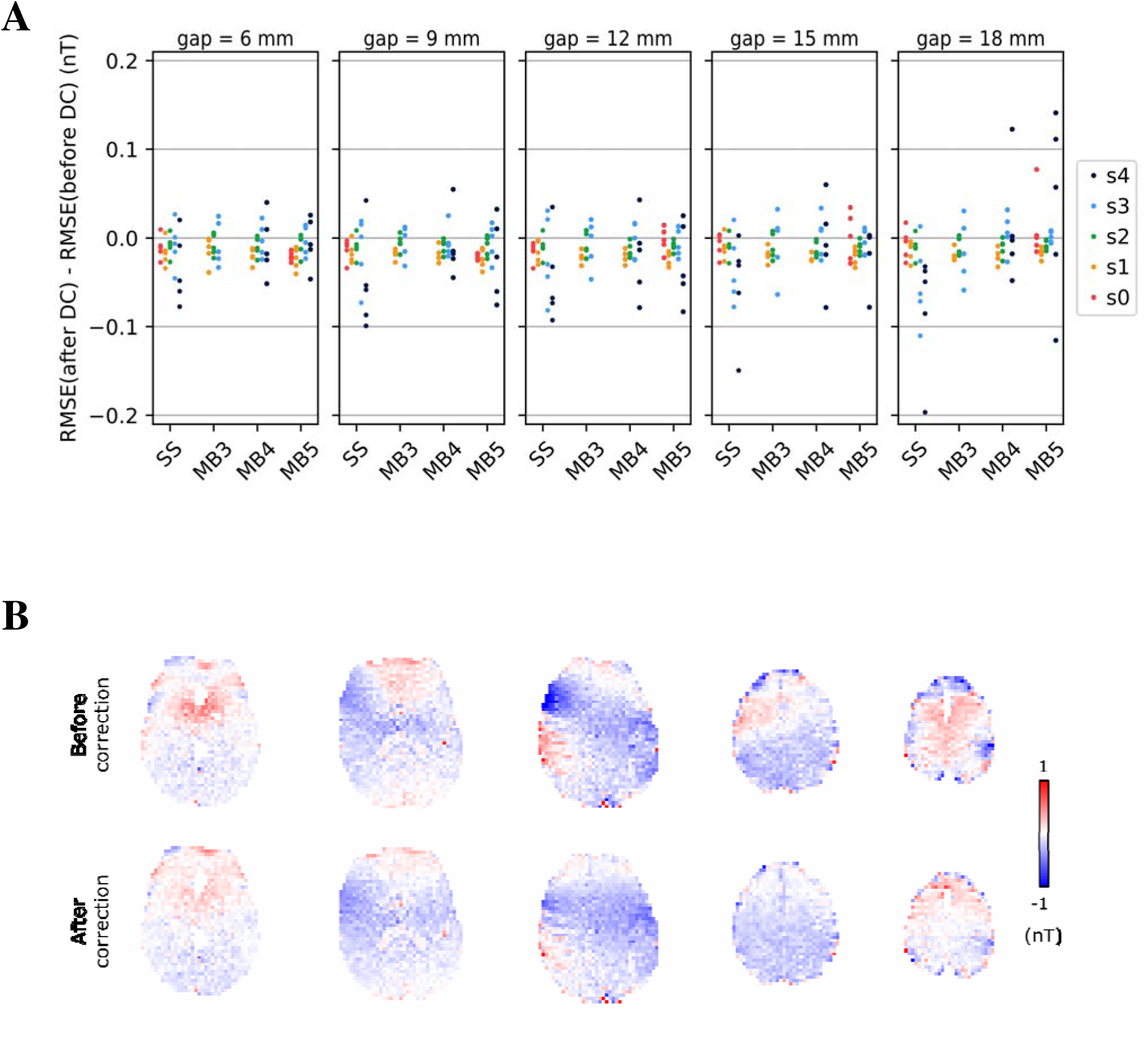

**Supplementary Figure S2.**
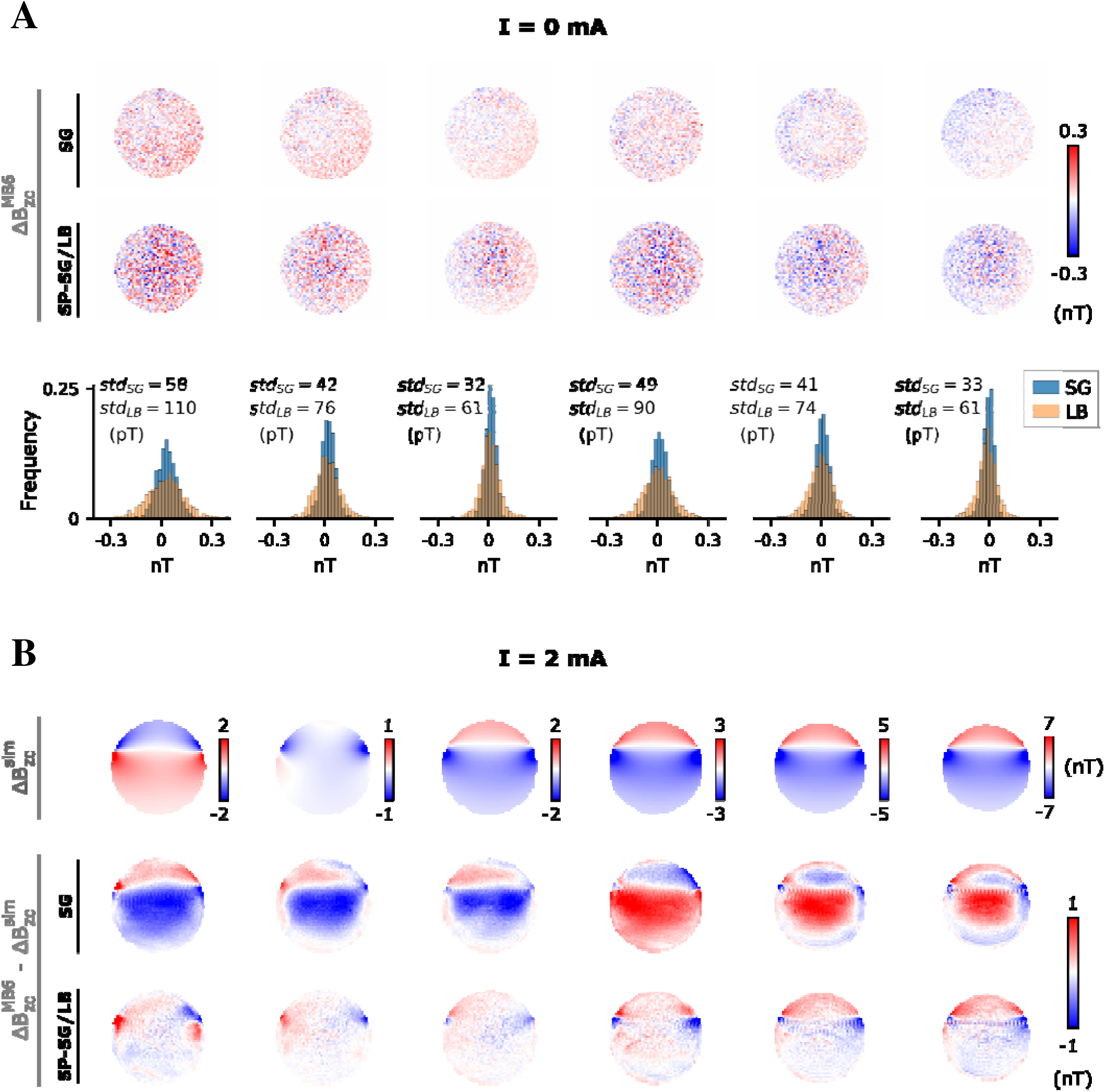

**Supplementary Figure S3.**
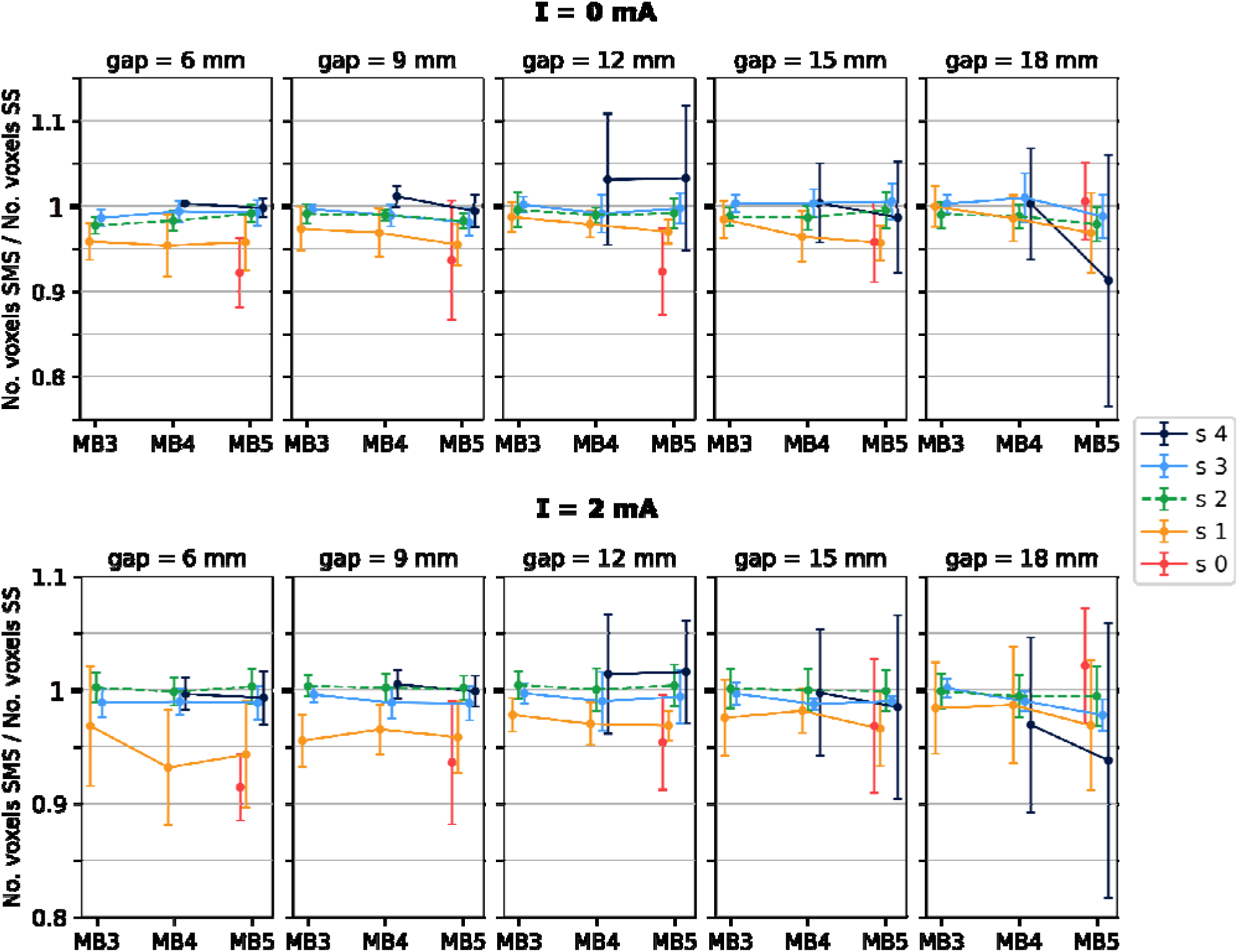

