## Supplementary_Material for "Increased brain coverage and efficiency when measuring current-induced magnetic fields by use of simultaneous multi-slice echo-planar MRI"

### Supporting Information

#### Supporting Material S1: Masking of $\Delta \mathrm{B}_{\mathrm{z,c}}$

The $\Delta \mathrm{B}_{\mathrm{z,c}}$ measurements were masked to exclude regions with low SNR caused by strong susceptibility gradients, low coil sensitivities, liquid motion or undesired coherence pathways due to insufficient spoiling of signal from long T2* tissues like the cerebrospinal fluid [1]. The final masks resulted from a multi-step procedure: An initial mask was obtained by setting a threshold of 7.5 on a smoothed version of the temporal SNR map of the magnitude images at the latest echo. Smoothing was performed with an isotropic 2D Gaussian kernel with a width of one voxel to mitigate the appearance of holes and disconnected components in the masks. The masked $\Delta \mathrm{B}_{\mathrm{z,c}}$ images were then corrected for geometric distortions caused by $\mathrm{B}_0$ inhomogeneities using field maps estimated from the phase images at the two echo times, as described in Supporting Material S2. Regions where the estimated susceptibility gradients along the phase-encoding direction were greater than 50 $\mu\mathrm{T/m}$ were further excluded due to the severity of the associated distortions. Disconnected components with less than 25 voxels resulting from this procedure were also removed. Finally, the intersections of the masks from the different acquisitions were calculated using the logical "and" operation to maximize consistency between the regions included in the comparison of the single- and multi-slice results.

#### Supporting Material S2: Distortion correction of $\Delta \mathrm{B}_{\mathrm{z,c}}$

##### Theory

The effects of $\mathrm{B}_0$ inhomogeneity during signal readout depend on the strength of the readout gradient. In 2D EPI, the phase-encoding (PE) blips can be modeled as a weak continuous readout gradient, which is the time average of the blips' moment during a complete traversal of k-space. Because the PE "readout" gradient is much weaker than the frequency-encoding one, the geometric distortions can be assumed to a good approximation to occur only in the PE direction. Thus, for simplicity, these effects will be derived for a 1D imaging experiment. Neglecting relaxation effects, the acquired k-space signal is given by

$$S\left(k(t)\right) \propto \int_{\mathbb{R}} M_{\perp}(y) e^{-i\Delta\phi(y,t)} e^{-i2\pi\cdot k(t)\cdot y} dy \ , \tag{1}$$

where $k(t) \equiv \frac{\gamma}{2\pi}\int_0^t G_{im}(t')dt'$ is the location in k-space sampled at time $t$ after excitation, $G_{im}$ is the imaging gradient, $y$ is the coordinate in object space, $M_{\perp}$ is the complex transversal magnetization, and the complex exponentials $e^{-i\Delta\phi(y,t)}$ and $e^{-i2\pi\cdot k(t)\cdot y}$ are the phases accrued due to $\mathrm{B}_0$ inhomogeneities and applied imaging gradients, respectively. Assuming a static $\mathrm{B}_0$ inhomogeneity $\Delta B(y)$, one can write $\Delta\phi(y,t) = \gamma\cdot\Delta B(y)\cdot t$. A prewinder gradient is applied before the main readout gradient, allowing the center of k-space to be sampled at the echo time $T_E$. Thus, a new time variable $\tau = t - T_E$ is introduced, which is directly proportional to the k-space coordinate $k$ for a constant readout gradient (in EPI, we consider the time average of the gradient blips, $\bar{G}_{im}$): $k(\tau) = \frac{\gamma}{2\pi}\cdot\bar{G}_{im}\cdot\tau$. Incorporating this into equation 1 results in

$$S\left(k(\tau)\right) \propto \int_{\mathbb{R}} M_{\perp}(y) e^{-i\gamma\cdot\Delta B(y)\cdot T_E} e^{-i\gamma\cdot\bar{G}_{im}\cdot\left(y+\frac{\Delta B(y)}{\bar{G}_{im}}\right)\cdot\tau} dy \ . \tag{2}$$

The following change of variable can be made:

$$y' = y + \frac{\Delta B(y)}{\bar{G}_{im}} \tag{3a}$$

$$\frac{dy'}{dy} = 1 + \frac{G(y)}{\bar{G}_{im}} \equiv Q(y) \ , \tag{3b}$$

where $y'$ can be interpreted as a coordinate in the new (distorted) space, $G(y) = \frac{d\Delta B(y)}{dy}$ is the spatial gradient of the field inhomogeneity in the original (undistorted) space, and $Q(y)$ is defined as the local spatial stretching. In a first order approximation, $\Delta B(y)$ around the location $y_0$ can be written as

$$\Delta B(y) \approx \Delta B(y_0) + G(y_0)\cdot(y - y_0) \ . \tag{4}$$

Combining equations 3a, 3b and 4 results in

$$y = \frac{1}{Q(y_0)}\left[y' - \frac{1}{\bar{G}_{im}}\Big(\Delta B(y_0) - G(y_0)\cdot y_0\Big)\right] \equiv f(y') \ , \tag{5}$$

and equation 2 becomes

$$S\left(k(\tau)\right) \propto \int_{\mathbb{R}} \frac{M_{\perp}\big(f(y')\big)}{Q\big(f(y')\big)} e^{-i\gamma\cdot\Delta B(f(y'))\cdot T_E} e^{-i\gamma\cdot\bar{G}_{im}\cdot y'\cdot\tau} dy' \ . \tag{6}$$

A simple inverse Fourier Transform of the k-space signal in equation 6 results in a distorted image $I(y')$ with voxel displacements, magnitude scaling and an additional phase term. The term $\Delta B\left(f(y')\right)$ can be expanded with the help of equations 4 and 5, resulting in

$$\Delta B'(y') \equiv \Delta B\left(f(y')\right) \approx \frac{\Delta B(y_0) + G(y_0)\cdot(y' - y_0)}{Q(y_0)} \ . \tag{7}$$

In MRCDI, two components of $\Delta B$ should be distinguished: The tiny current-induced magnetic fields, $\Delta B_{z,c}$, and all other static $\mathrm{B}_0$ inhomogeneities, $\Delta B_*$. As the shifts caused by $\Delta B_{z,c}$ are almost negligible compared to those due to the much larger $\Delta B_*$, the coordinate $y_0$ is approximately mapped to $y_0' = y_0 + \Delta B_*(y_0)/\bar{G}_{im}$ (equation 3a). A first order approximation of $\Delta B'(y')$ around this point is given below:

$$\Delta B'(y') \approx \frac{\left(\Delta B + \frac{G}{\bar{G}_{im}}\Delta B_*\right)(y_0) + G(y_0)\cdot(y' - y_0')}{Q(y_0)} \tag{8}$$

**Current-induced magnetic fields**

Inverting the polarity of the applied currents only affects the sign of the $\Delta B_{z,c}$ component of $\Delta B'$. Therefore, at $y' = y_0'$, it is given by

$$\Delta B'_{\pm}(y')|_{y'=y_0'} \approx \left(\frac{\Delta B_* \pm \Delta B_{z,c} + \frac{G_* \pm G_{z,c}}{\bar{G}_{im}}\Delta B_*}{1 + \frac{G_* \pm G_{z,c}}{\bar{G}_{im}}}\right)(y_0)\ . \tag{9}$$

Subtracting the phase of two images acquired with opposite current polarity thus gives

$$(\Delta\phi_- - \Delta\phi_+)(y')|_{y'=y_0'} = 2\gamma\cdot\left(\frac{1 + G_*/\bar{G}_{im}}{1 + 2G_*/\bar{G}_{im}}\Delta B_{z,c}\right)(y_0)\cdot T_E\ . \tag{10}$$

When the gradient of the field inhomogeneity is much smaller than the average imaging gradient ($G_* \ll \bar{G}_{im}$), a first order approximation can be made:

$$(\Delta\phi_- - \Delta\phi_+)(y')|_{y'=y_0'} \approx 2\gamma\cdot\frac{\Delta B_{z,c}}{Q_*}(y_0)\cdot T_E\ , \tag{11}$$

with $Q_* = 1 + G_*/\bar{G}_{im}$. The ratio $T_E/Q_*$ can be interpreted as the echo time shifting effect described in [2]. The current-induced magnetic fields computed in the distorted space, $\Delta B'_{z,c}$, differ from the true magnetic fields, $\Delta B_{z,c}$, by both voxel displacements and intensity scaling:

$$\Delta B'_{z,c}(y_0') \equiv \frac{(\Delta\phi_- - \Delta\phi_+)(y_0')}{2\gamma\cdot T_E} \approx \frac{\Delta B_{z,c}}{Q_*}(y_0) \tag{12}$$

**EPI-based field maps**

When no electrical currents are applied, equation 8 can be simplified to

$$\Delta B'_*(y') \approx \Delta B_*(y_0) + \frac{G_*}{Q_*}(y_0)\cdot(y' - y_0')\ . \tag{13}$$

Thus, the phase difference at $y' = y_0'$ between two images acquired at different echo times, with otherwise identical conditions, is given by

$$\left(\Delta\phi_{T_{E,2}} - \Delta\phi_{T_{E,1}}\right)(y')|_{y'=y_0'} \approx \gamma\cdot\Delta B_*(y_0)\cdot\Delta T_E\ , \tag{14}$$

with $\Delta T_E = T_{E,2} - T_{E,1}$ the echo time difference. The static field inhomogeneities computed in the distorted space, $\Delta B'_*$, differ from the true undistorted ones, $\Delta B_*$, by voxel displacements only:

$$\Delta B'_*(y'_0) \equiv \frac{\left(\Delta\phi_{T_{E,2}} - \Delta\phi_{T_{E,1}}\right)(y'_0)}{\gamma \cdot \Delta T_E} \approx \Delta B_*(y_0) \tag{15}$$

#### Implementation

A complete distortion correction pipeline including the computation of EPI-based field maps, as well as displacement and intensity correction routines, was implemented in Python version 3.8.5 [3]. Briefly, the dual-echo EPI sequence yielded two images at different echo times for each excitation pulse. The resulting time series of images was averaged at each echo time to improve the SNR. A phase difference image was then computed and phase wraps were removed using PRELUDE from the FMRIB Software Library (FSL 6.0.4, https://fsl.fmrib.ox.ac.uk/fsl/). Under the assumption that the $\mathrm{B_0}$ inhomogeneities were constant in time, an estimate of the field map in the distorted space was obtained by dividing the phase difference image by the echo time difference. Light smoothing was then applied using an isotropic 2D Gaussian kernel with a width of one voxel in both directions to further reduce the noise. A field map in Hz differs from the map of voxel displacements by the scaling factor $\mathrm{ETL} \cdot \mathrm{t_{esp}}$, where $\mathrm{ETL}$ is the EPI echo train length and $\mathrm{t_{esp}}$ is the echo spacing [4]. A voxel shift map in the undistorted space was then obtained from a piece-wise linear interpolation, along the PE direction, of the corresponding distorted version [5]. A field map in the undistorted space was recovered based on the proportionality described above, and its gradient along the PE direction was computed to characterize the intensity correction factor Q defined in equation 3b. The $\Delta\mathrm{B_{z,c}}$ images were finally corrected for the two main effects of $\mathrm{B_0}$ inhomogeneity: Voxel displacements and intensity modulation.

The implemented distortion correction pipeline decreased the difference between the predicted and measured fields for most of the tested cases (see Figure S1 below), validating its usage for the results presented above in the main part of the paper. Wilcoxon signed-rank tests were performed on data pooled per slice number (s0 to s4) to reach sufficient power per test and confirmed statistically significant reductions ($\mathrm{p} < 0.001$) for all five slice numbers.

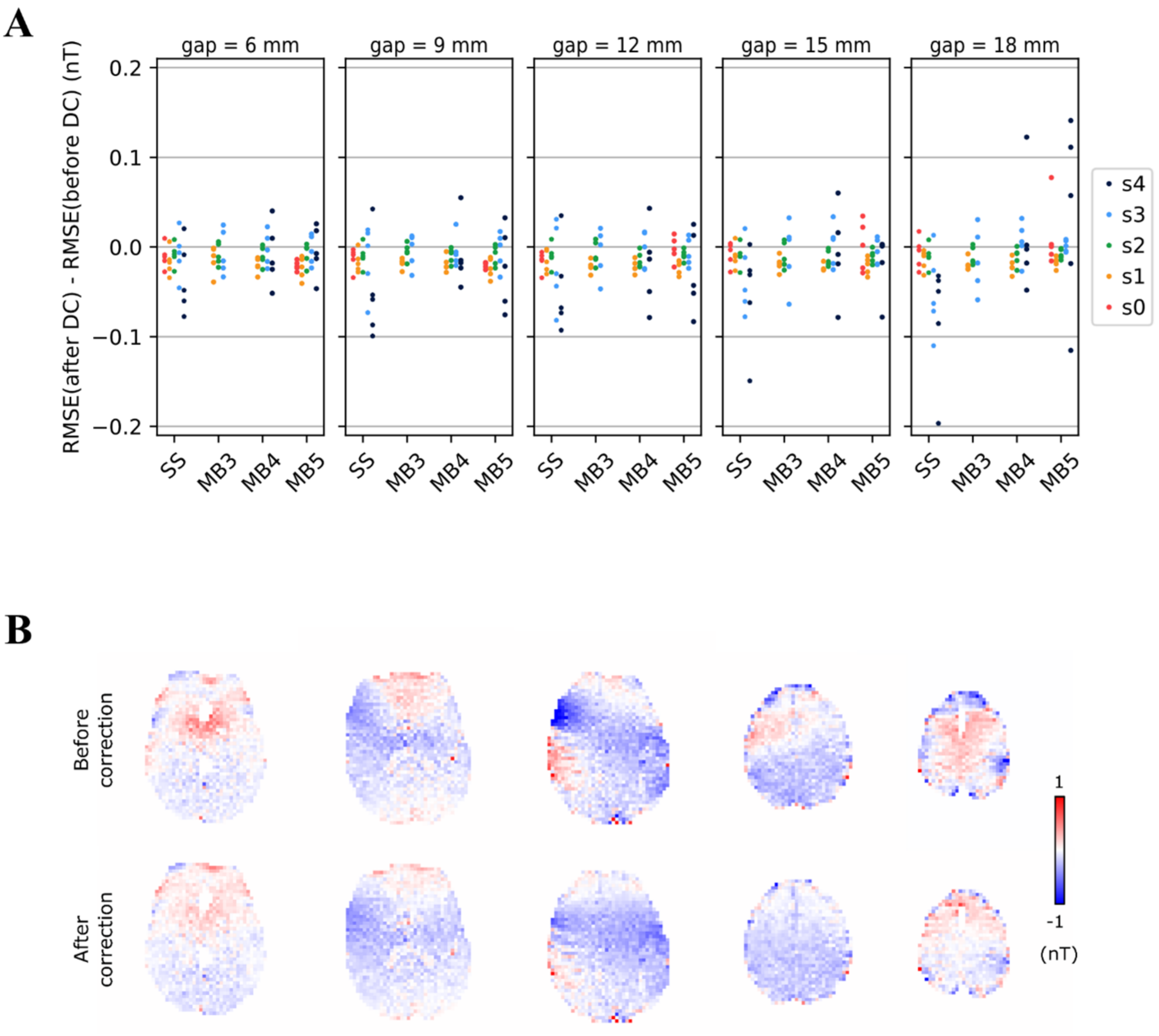


**Supporting Figure S1. A)** Change in the root mean square error (RMSE) between measured and simulated magnetic fields after application of the implemented distortion correction (DC) pipeline to the measured fields. For each slice (s0 to s4), each data point corresponds to one of the five scanned subjects. The distortion correction reduced the error between measurement and simulation in most cases. **B)** Example of the correction of the current-induced magnetic fields ($\Delta B_{z,c}$) for the effects of main field ($B_0$) inhomogeneity. The images depict the differences between the measured and simulated $\Delta B_{z,c}$ before and after distortion correction of the measured $\Delta B_{z,c}$ with the implemented pipeline, for five example slices. Because the voxel displacements were small (less than two voxels), the most visible consequence of $B_0$ inhomogeneity is the modulation of the $\Delta B_{z,c}$ intensity. Note the increased similarity between the simulated and measured $\Delta B_{z,c}$ after distortion correction, as reflected by the substantial attenuation of the patterns seen in the difference images.

### Supporting Material S3: Additional Figures

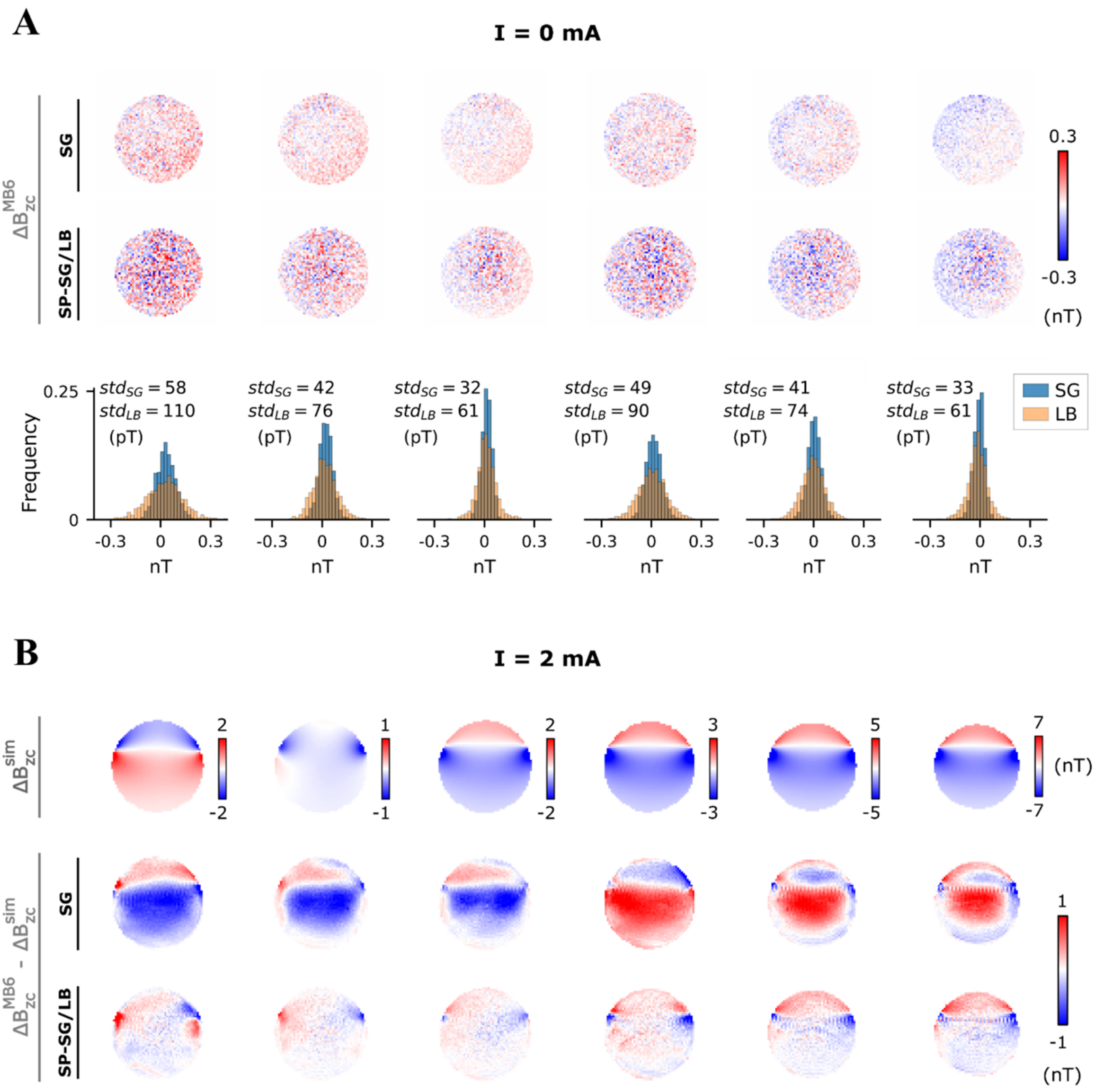


**Supporting Figure S2.** Comparison between two approaches for separating the simultaneously acquired slices: Slice-GRAPPA (SG) and Split Slice GRAPPA/"LeakBlock" (SP-SG/LB). Examples are given for a multiband factor of 6 and a 6 mm interslice gap. **A)** $\Delta B_{z,c}$ measurements without current injection. The noise floor images obtained with each slice-separation technique are shown in the first two rows, with the associated histograms and standard deviations summarized in the third row. In this current-free scenario, where no time-varying phase changes were induced by the application of electric currents, the SG technique showed a superior performance, with the spatial standard deviation of $\Delta B_{z,c}$ being consistently lower in all slices. **B)** $\Delta B_{z,c}$ measurements with current injection. The magnetic fields simulated according to the Biot-Savart Law are shown in the first row, while the differences between the measured and simulated fields (residuals) are shown in the second and third rows for the different slice-separation approaches. Upon application of time-varying electric currents, the SP-SG/LB approach provided a much more robust reconstruction, as judged by the substantially lower residuals.

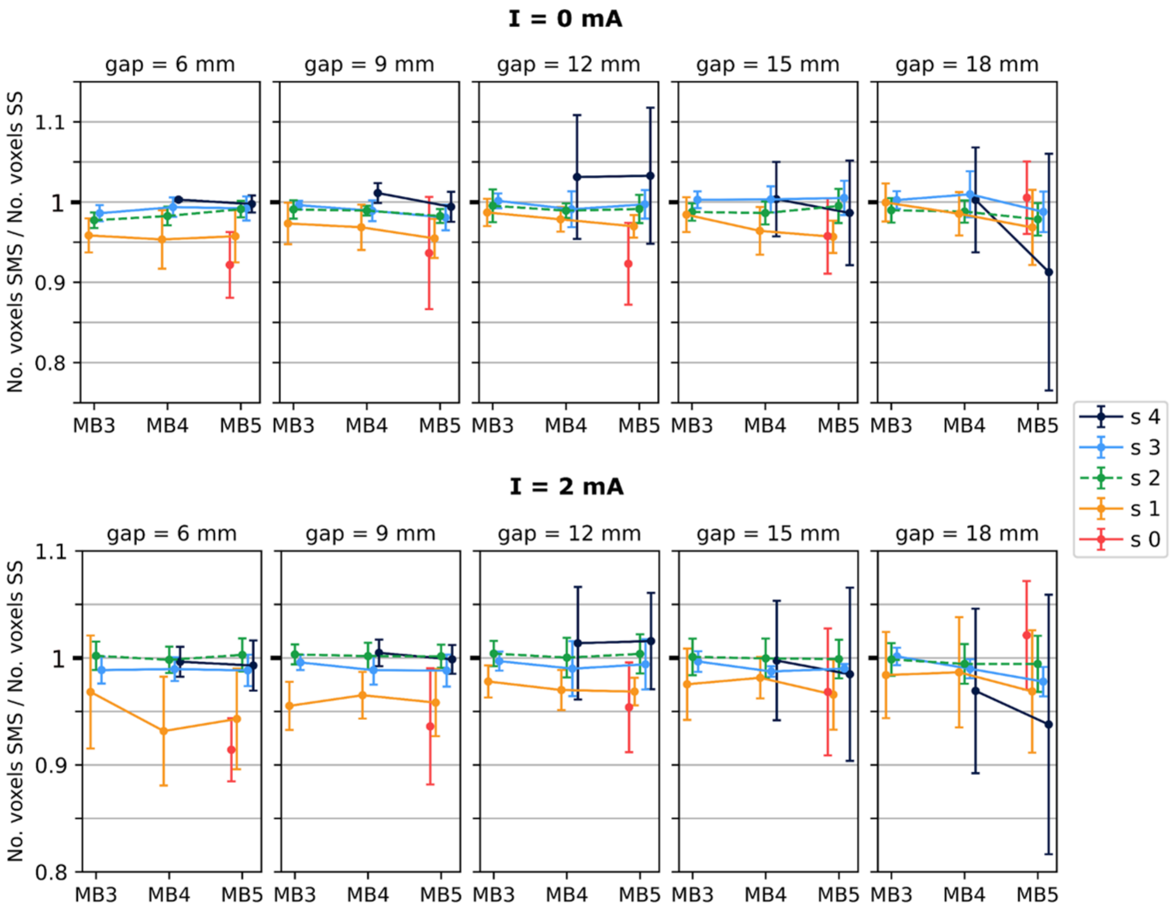


**Supporting Figure S3.** Quantification of the number of voxels preserved after masking based on the temporal signal-to-noise ratio of the magnitude images. The results are summarized as the number of voxels preserved in each slice of a simultaneous multi-slice (SMS) acquisition, normalized to those of the corresponding single-slice (SS) acquisition. Each data point and error bar represent the mean and standard deviation across five subjects. The top and bottom plots refer to the experiments without and with electric currents, respectively. On average across subjects, the number of voxels preserved did not differ by more than 10% between the two acquisition types, and was below 5% in most cases.
